# Working memory recovery in adolescents with concussion: Longitudinal fMRI study

**DOI:** 10.1101/2024.05.11.24307147

**Authors:** A. Manelis, J.P. Lima Santos, S.J. Suss, C.A. Perry, R.W. Hickey, M.W. Collins, A.P. Kontos, A. Versace

## Abstract

**Background:** Understanding behavioral and neural underpinnings of post-concussion recovery of working memory function is critically important for improving clinical outcomes and adequately planning return-to-activity decisions. Previous studies provided inconsistent results due to small sample sizes and the use of a mixed population of participants who were at different post-injury time points.

**Methods:** To better understand behavioral and neural correlates of working memory recover during the first 6 months post-concussion, we used functional magnetic resonance imaging (fMRI) to scan 45 concussed adolescents [CONC] at baseline (<10 days post-concussion) and again at 6 months post-concussion. Healthy control [HC] adolescents without a history of concussion were scanned once. During the scans, participants performed 1-back (easy) and 2-back (difficult) working memory tasks with the letters as the stimuli and angry, happy, neutral, and sad faces as distractors.

**Results:** By the 6-month follow-up, all affected adolescents were asymptomatic and cleared to return-to-activity. Working memory function recovery was reflected by faster and more accurate performance at 6 months vs. baseline (p-values<0.05). It was also characterized by significant difficulty-related activation increases in the left inferior frontal gyrus (LIFG) and the left orbitofrontal cortex (LOFC) at 6 months vs. baseline.

**Conclusion:** Post-concussion recovery is associated with significant performance improvements in speed and accuracy, as well as normalization of brain responses in the LIFG and LOFC during the n-back task. The observed patterns of LOFC activation might reflect compensatory strategies to distribute neural processing and reduce neural fatigue post-concussion.

## 1 INTRODUCTION

The lifetime prevalence of concussion among adolescents is rising and increased from 19.5% in 2016 to 24.6% in 2020 [1]. Post-concussion recovery usually takes around four weeks, but in some cases may take longer [2]. Longitudinal studies comparing cognitive and brain function changes during recovery may shed light on the mechanisms underlying post-concussion impairments and recovery.

Post-concussion symptoms in adolescents impact their cognitive and social function [3,4] in addition to eliciting movement-related dizziness, nausea, vision alteration, excessive fatigue, and other physiological symptoms [5–7]. Working memory, which is the ability to update and manipulate information online [8,9], is one of the cognitive domains frequently affected by concussion [10,11]. Performance on the working memory tasks relies on the prefrontal, cingulate, and posterior parietal cortical regions that increase in activation when the task becomes more difficult [8,12–16]. These regions are actively developing in adolescents [17–20], so if concussion affects these regions, development of cognitive function may also be impacted.

Working memory is often assessed using n-back tasks in which participants are presented with a sequence of stimuli and have to indicate whether the current stimulus is the same as the stimulus presented one or more items before [12,15,16]. Previous cross-sectional neuroimaging studies have reported inconsistent findings regarding the impact of concussion on working memory task performance and corresponding brain activation patterns. For example, several studies found no differences in behavioral performance on working memory tasks between concussed and control individuals at one month [21], 7.5 months [22], or 2.5 years [23] post-concussion. However, other studies found reduced working memory task performance in the acute/subacute post-concussion period [16,24] as well as three months after injury [25]. Working memory task performance was associated with lower prefrontal cortical and parietal activation [24,25] in concussed vs. control adolescents. More symptomatic participants showed greater differences in retrosplenial cortex activation for the 1-back vs. 2-back condition than their less symptomatic counterparts within the first 10 days of injury [16]. Brain activation in the temporal and frontal brain regions was greater in concussed adolescents during the 2-back condition at 7.5 months [22]. There were no significant differences in working memory-related brain activation between concussed and control adolescents 2.5 years after injury [23].

The findings cited above suggest that cross-sectional studies are not ideal for capturing changes across post-concussion recovery. Therefore, conducting longitudinal studies that compare working memory performance immediately after concussion with that after full clinical recovery is critically important. Only a small number of studies have examined post-concussion recovery longitudinally. One study scanned youth athletes three times: within 72 hours, at two weeks, and two months post-concussion [26]. Despite comparable behavioral performance at all time points, Detwiller et al. reported greater activation in bilateral dorsolateral prefrontal and inferior parietal cortices in concussed participants relative to controls for 2-back vs. 1-back conditions at baseline and two weeks post-concussion[26]. The other study scanned adults one month after a concussion and six weeks after the first scan. They found that affected individuals were not able to adequately increase their brain activation for difficult vs. easy working memory tasks one month after injury. This ability, however, improved at the six-week follow-up [27]. A third study examined behavioral and brain alterations after one month of post-concussion recovery and found that performance on a difficult working memory task became more accurate and elicited greater prefrontal activation in concussed adolescents [11].

The inconsistent results reported by the previous cross-sectional and longitudinal studies could be explained by small sample sizes and inclusion of a mixed population of participants who were not fully recovered with those who were recovered after concussion. The goal of our study was to examine post-concussion recovery of working memory in affected adolescents by comparing their behavioral performance and brain activation during the acute/subacute period (<10 days after concussion) with that at six months after injury. Based on the extant literature briefly reviewed above, we hypothesized that post-concussion recovery of working memory would be characterized by activation changes in the prefrontal, parietal, and PCC regions during performance on difficult vs. easy working memory tasks. We also hypothesized that behavioral performance (i.e., accuracy and reaction time (RT)) would improve across recovery and become similar to that of HC.

## 2 METHODS

### 2.1 Participants

The study was approved by the University of Pittsburgh Institutional Review Board (IRB number STUDY19030360). Written informed assent was obtained from all participants, and written informed consent was obtained from all caregivers. All participants were adolescents between 12-17 years of age. Adolescents with concussion (CONCUS) were diagnosed using current consensus criteria (McCrory et al., 2017) within 1-10 days after injury. Those who lost consciousness for more than 5 minutes at the time of the injury were excluded from the study. Healthy control adolescents (HC) had no history of concussion and were age- and sex-matched to those with concussion. Exclusion criteria for all adolescents included IQ below 70, MRI contraindications, systemic medical, neurologic, psychotic spectrum, and neurodevelopmental disorders, orthopedic injury within the past month, alcohol or illicit substance use/dependency within the past three months, intoxication, or presence of illicit drugs in a urine test on the day of the scan and left/mixed handedness.

CONC were scanned twice – at baseline during the subacute phase of concussion recovery (1-10 days after concussion), and 6 months after concussion. Only the individuals who attended both scans and had usable data were included in the longitudinal data analysis. The HC were scanned one time. A total of 77 adolescents (45 CONC and 32 HC) met eligibility criteria, had usable fMRI data, and therefore, were included in the behavioral and neuroimaging data analyses. Of these individuals, cross-sectional findings have already been reported for 26 CONC and 27 HC [16].

### 2.2 Clinical Assessments

The presence of psychiatric disorders and/or current use of psychotropic medications was examined using the International Neuropsychiatric Interview for children and adolescents (MINI-KID) [28]. Concussion symptoms were measured using Post-Concussion Symptom Scale (PCSS) [29], and Vestibular/Ocular-Motor Screening (VOMS) [6] that were administered by a trained clinician. Higher scores on all three assessments indicated greater severity. Considering that in our previous cross-sectional study of subacute concussion only the VOMS scores were associated with the changes in brain activation during the n-back task, we used this measure as a covariate in the behavioral neuroimaging data analysis in this longitudinal study. All relevant demographics and medical history as well as the mechanism of concussion are reported in Table 1.

**Table 1.** Demographics and clinical data for adolescents with concussion (i.e., Concussed) and healthy control adolescents (i.e., Controls)

|  | **Concussed** | **Controls** | **statistics** |
| --- | --- | --- | --- |
| N | 45 | 32 |  |
| Number of females (%) | 16 (35%) | 16 (50%) | $\chi^2 (1) = 0.4, p = 0.5$ |
| Race<br>• Number of Caucasian (%)<br>• Number of Black (%)<br>• Number of Unknown or more than one race (%) | • 38 (85%)<br>• 7 (13%)<br>• 0 | • 34 (78 %)<br>• 5 (19%)<br>• 3 (4%) | $\chi^2 (3) = 4.4, p = 0.2$ |
| Mean age in years (SD)<br>[min-max] | 15.9 (1.3)<br>[13-17.9] | 15.5 (1.2)<br>[13-17.4] | $t(75) = 1.3, p = 0.2$ |
| Mean IQ (SD)<br>[min, max] | 104.3 (8.4)<br>[83.0-122.0] | 107.9(8.0)<br>[92.0-124.0] | $t(77) = -1.8, p = 0.07$ |
| Mean VOMS scores total (SD)<br>[min, max] | 43.9(43.4)<br>[0.0-193.0] | - | na |
| Mean PCSS scores total (SD)<br>[min, max] | 28.9(21.3)<br>[0.0-79.0] | - | na |
| Mean number of days between injury and scan (SD)<br>[min, max] | 3.1(2.3)<br>[0-9] | - | na |
| Mean number of days between baseline scan and 6-month scan (SD)<br>[min, max] | 190.3(13)<br>[168-209] | - | na |
| Mean number of days between injury and clearance for return to play (SD)<br>[min, max] | 30.2 (33.9)<br>[2 – 175] | - | na |
| Mean number of days between the clearance date and 6-month scan (SD)<br>[min, max] | 159.9(40)<br>[25-200] | - | na |

### 2.3 Experimental paradigm

The details of the n-back task used in this study to measure working memory are described elsewhere [16]. In short, participants were presented with lowercase and capital letters one at a time. In the 1-back condition, they had to indicate whether the letter presented on the screen matched the one shown in the previous trial. In the 2-back condition, participants had to indicate whether the letter presented on the screen matched the letter presented two trials ago. On each trial, the letters were presented with either angry, happy, neutral, or sad faces located on the right and left sides of the letter (Figure 1). The faces were taken from the NimStim database [30] and served as emotional distractors [31]. Participants performed three 5-minute runs consisting of eight 30-second blocks of trials (1-back and 2-back tasks with 4 emotional face distractor conditions) presented in random order and preceded by a 4.5-second instruction screen. Participants had to respond as quickly and accurately as possible by pressing the response button with the index finger of one hand for the letters that were in the 1- or 2-back position and with the other hand for all other stimuli. We counterbalanced hand assignment across participants.

**Figure 1.**
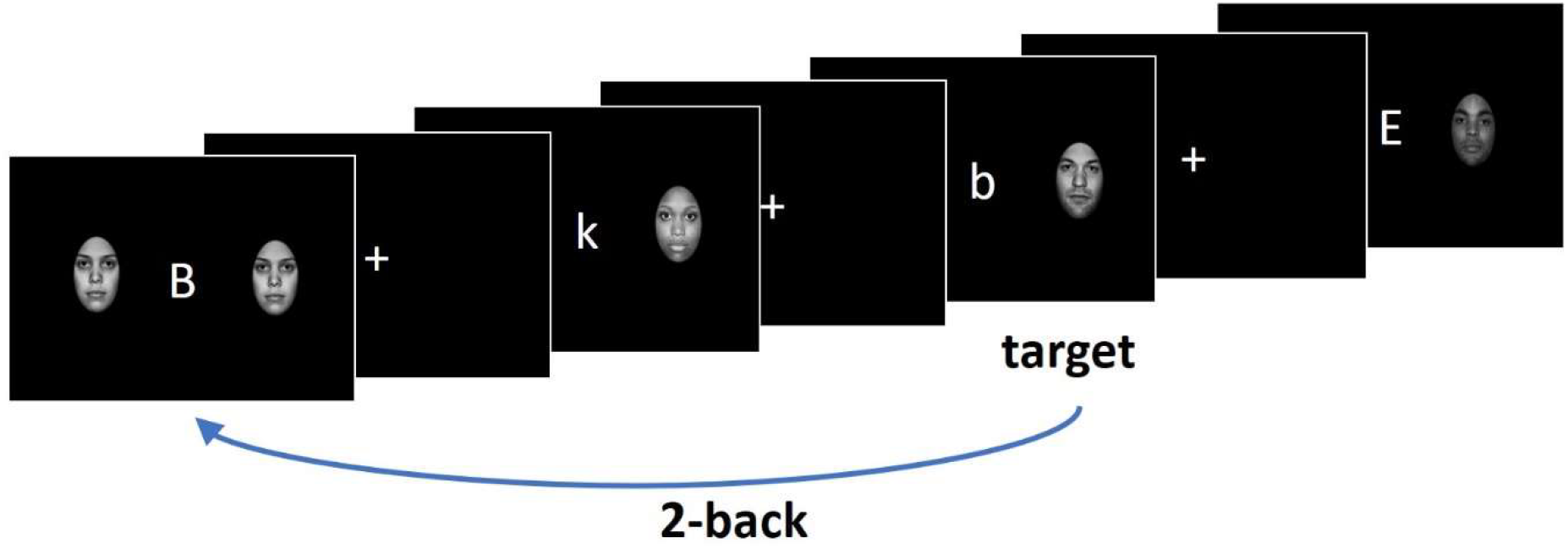
The n-back task paradigm (the figure is adopted from [16]).

**Figure 2.**
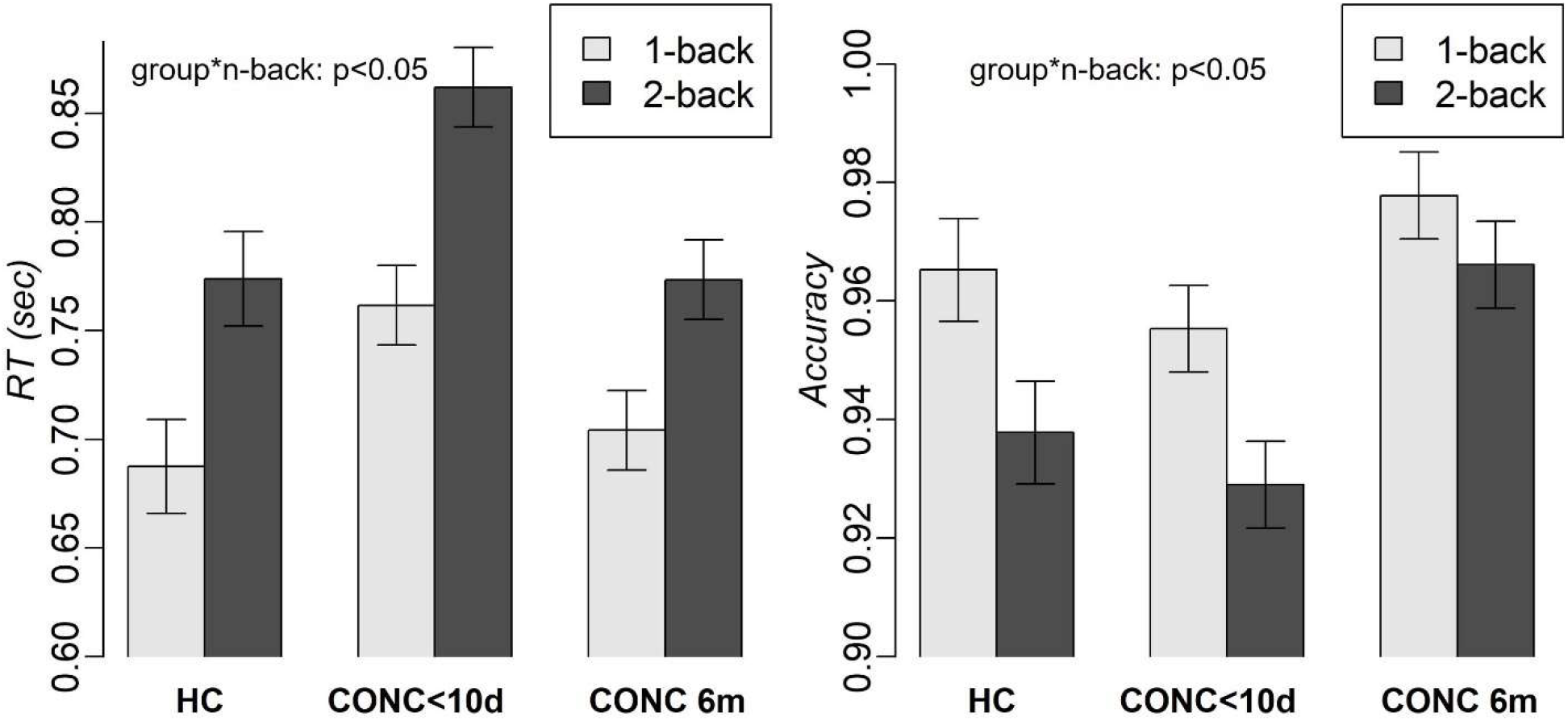
RT and accuracy in HC and adolescents with concussion during the first 10 post injury and post-recovery (6 months post injury) periods.

### 2.4 Behavioral data analysis

We used three-way mixed effects linear models (‘*lme4’* package in R (Bates et al., 2015)) with group (CONC 1, CONC 2, HC), n-back condition (1- and 2-back), and distractor emotions (happy, angry, sad, neutral) as independent variables to explain RT and accuracy (in the separate models) using Type III Analysis of Variance with Satterthwaite’s method. Participants were treated as a random factor and their age, sex and VOMS scores at baseline served as covariates. When appropriate, the contrasts and means were estimated from the mixed effects models (‘*modebased’* package in R (Makowski et al., 2020)). Tukey’s Honestly Significant Difference (HSD) was used to correct for multiple comparisons.

### 2.5 fMRI data acquisition

Functional MRI data were acquired at the University of Pittsburgh using a Siemens PRISMA 3T MR system and a 32-channel RF head coil. The MPRAGE sequence was used to acquire a high-resolution structural image (voxel size=1×1×1mm^3^, TR= 2400ms, FOV=256mm, flip angle=8°, 176 slices). A gradient-echo, echo-planar sequence was used to collect functional MRI data (360 volumes per run; voxel size: 2×2×2 mm^3^, TR=800ms, echo time = 30ms, field of view = 210mm, flip angle = 52°, 72 slices, multiband acceleration factor=8). Two spin echo images with the anterior-to-posterior and posterior-to-anterior phase encoding directions (voxel size=2×2×2mm^3^, TR=8000ms, TE=66.00ms, FOV=210mm, flip angle=90°, 72 slices) were collected to allow for geometric distortion correction of fMRI data.

### 2.6 fMRI data preprocessing

DICOM files were converted to NIFTI using the *dcm2niix* tool (v1.0.20180826 BETA GCC5.4.0)[32]. Non-brain tissues were removed using the optiBET script [33]. Motion correction was performed using MCFLIRT [34]. Spatial smoothing with a Gaussian kernel of full-width at half-maximum = 6 mm was applied. Susceptibility distortion in the BOLD images was corrected using *topup* in FSL6.0.3 (www.fmrib.ox.ac.uk/fsl). BOLD images were transformed to MNI space by first registering BOLD images to the high-resolution structural (MPRAGE) images using FLIRT (FMRIB’s Linear Image Registration Tool) [34,35] with Boundary-Based Registration (BBR), and then registering the high-resolution images to the MNI152 T1-2mm template using FNIRT (FMRIB’s Non-linear Image Registration Tool) [36] with nine degrees of freedom (DOF). The two resulting transformations were concatenated and applied to the original BOLD. The quality of transformation was examined.

Motion artifacts were removed from BOLD images using ICA-AROMA [37]. High-resolution structural images were segmented to white matter, grey matter, and cerebral-spinal fluid (CSF) using the *fsl_anat* script (http://fsl.fmrib.ox.ac.uk/fsl/fslwiki/fsl_anat) and the time courses associated with the white matter and CSF masks were extracted from the preprocessed functional data and regressed out during the analysis. After that, a high-pass filter (Gaussian-weighted least-squares straight line fitting, with sigma=56.25) was applied.

### 2.7 fMRI data subject-level and group-level analysis

#### 2.7.1 Subject-level analysis

A subject-level General Linear Model analysis was implemented using FEAT (FMRI Expert Analysis Tool, v6.0). A hemodynamic response function was modelled using a Gamma function. The model included regressors for instruction screens informing about 1- and 2-back blocks as well as for 1-back and 2-back blocks with happy, angry, fear and sad face distractors. The contrasts of interest included computing the 1-back vs. 2-back differences in brain activation across all face distractors as well as for each face distractor separately. The data from the available runs was averaged for each subject and session.

#### 2.7.2 Group-level analysis

The first group-level analysis included the CONC group only and compared the 1-back vs. 2-back differences in brain activation at baseline (post-concussion sub-acute phase) vs. the 1-back vs. 2-back differences at 6-month follow-up. This longitudinal analysis was conducted using the Sandwich Estimator (*swe*) approach [38] for nonparametric permutation inference conducted in the whole brain with Threshold-Free Cluster Enhancement correction (TFCE) [39], 1000 permutations, and FWE-corrected p-values< 0.05. The p-values were corrected for the 2 contrasts of interest (1-back>2-back and 2-back>1-back) using the Bonferroni method (p=0.05/2=0.025). Given that our previous cross-sectional study of concussed adolescents did not reveal the effect of distractor emotions on RT and accuracy [16], we decided to compare the two time points across all emotional distractors if the behavioral data analysis does not revealed the effect of emotions again. In contrast, we would examine the 2 timepoint x 4 emotions model if there was the effect of emotions on RT and accuracy.

The follow up analyses were conducted based on the percent signal changes extracted from the brain regions showing significant differences between sub-acute and 6-months scans. We used mixed-effects models (as described for the analysis of behavioral data) to compare brain activation in HC and CONC at two time points and to understand the relationship between brain activation and behavioral measures of working memory.

## 3 RESULTS

Unusable runs were excluded from both brain and behavior analyses. A total of 11 runs from sessions 1 and 2 were excluded. While the longitudinal analyses included only the CONC group (the HC group was scanned only once), we used the behavioral and brain data (extracted from the regions showing the changes in activation from baseline to 6 months) for HC during session 1 as a comparison.

### 3.1 Demographics and Behavioral

The CONC and HC groups did not differ in age, race, and biological sex profiles (Table 1). All adolescents with concussion were cleared to return to their activities at the 6 months scan (mean number of days between injury and clearance = 30.2 days, SD=33.9, range=2-175 days). There were on average 159.9 days (SD=40, range=25-200 days) between the date of clearance for activities and the follow-up scan at 6 months.

The mixed effects analysis with group, n-back condition, and distractor emotions as independent variables revealed a significant group—by—n-back interaction effect (F(2, 877.19)=3.3, p=0.04), and the main effects of n-back (F(1, 877.1)=265, p<0.001) and group (F(2, 134.96)=74.9, p<0.001) on RT. Slower RT was observed for 2-back vs. 1-back (t(895.02)=16.33, p<0.001) and for CONC subacute compared to both HC (1-back: t(84.28)=2.6, p=0.03; 2-back: t(84.28)=3.1, p=0.007) and CONC at 6-month (1-back: t(895.02)=6.8, p<0.001; 2-back: t(895.02)=10.4, p<0.001). RT did not significantly differ for HC vs. CONC at 6-month. We also observed the main effects of n-back (F(1, 876.32)=44.5, p<0.001) and group (F(2, 135.88)=31.6, p<0.001) on accuracy. Lower accuracy was observed for 2-back vs. 1-back (t(895.11)=-6.7, p<0.001) and for CONC subacute vs. CONC at 6-month (t(895.11)= -7.9, p<0.001). No significant effect of distractor emotions, age or biological sex was observed on either RT or accuracy.

### 3.2 Neuroimaging

Considering that we found no significant effect of distractor emotions on RT or accuracy, the neuroimaging data analysis was conducted across all emotions. The comparison of brain activation for 2-back—minus—1-back differences for subacute vs. 6-month follow-up scans in the CONC group revealed significant longitudinal changes in the left inferior frontal gyrus (LIFG; nvox=62, Z-max=4.9, [-40, 24, 16]) and the left orbitofrontal cortex (LOFC, nvox=104, Z-max=5.5, [-38, 34, -4]) (Figure3A).

**Figure 3.**
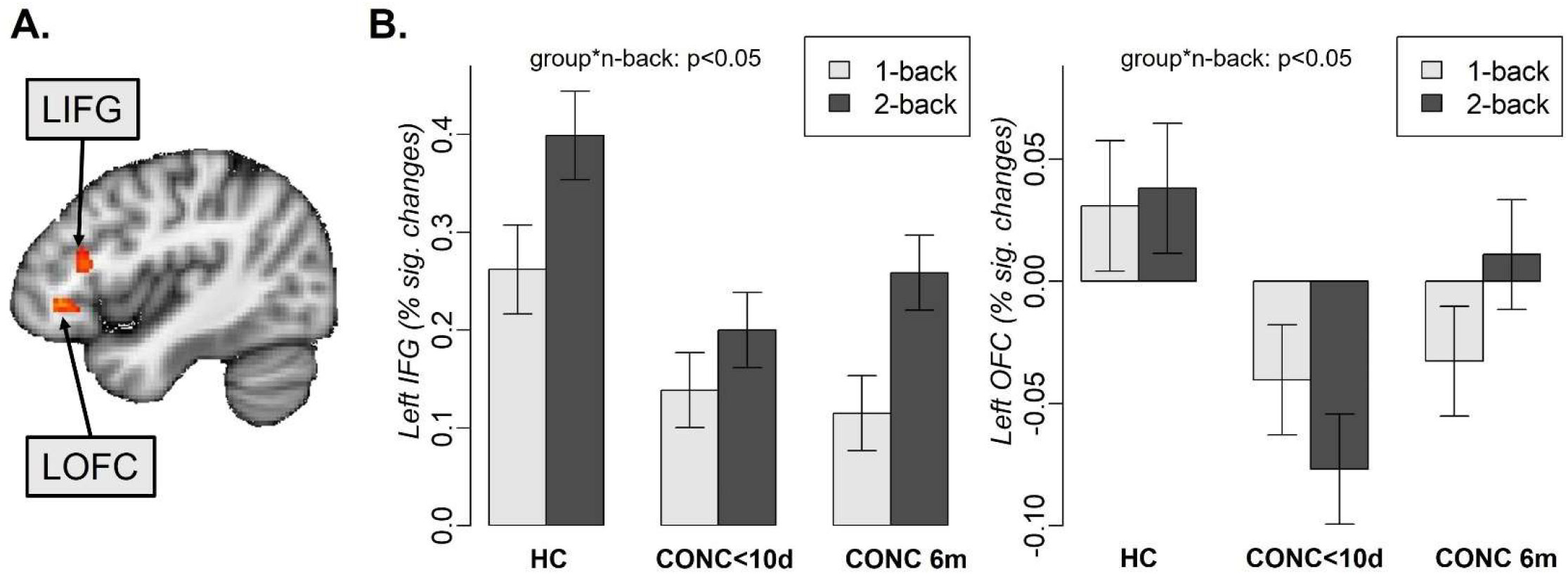
Left inferior frontal gyrus (LIFG) and left orbitofrontal (LOFC) activation during 1-back and 2-back tasks in healthy controls (HC), adolescents with concussion during the first 10 days (CONC<10d) and 6 months (CONC 6m) after concussion.

Further analyses were conducted on the percent signal changes extracted from these regions and included the CONC group at the two time points and HC at time point 1. To confirm that activation in the LIFG and the LOFC was not sensitive to distractor emotions, we included this variable in the mixed effects analysis described below. The mixed effects analysis with group, n-back condition, and distractor emotions as independent variables revealed the group-by-nback interaction (F(2, 877.16)=10.4, p<0.001), and the main effects of n-back (F(1, 877.16)=179.5, p<0.001) and group (F(2, 134.36)=5, p=0.008) on activation in the LIFG. HC had higher activation than CONC subacute (t(74.47)=2.75, p=0.02) and CONC at 6 month (t(74.47)=2.45, p=0.04). The same analysis in the LOFC revealed the group-by-nback interaction (F(2, 876.79)=8.5, p<0.001), and the main effects of group (F(2, 135.38)=14.3, p<0.001) (Figure 3B). CONC subacute had lower activation than HC (t(77.72)= -2.79, p=0.02) and CONC 6 month (t(77.72)= -4.9, p<0.001). There was no significant effect of distractor emotions, age, or biological sex on brain activation in either LIFG or LOFC activation.

The analysis of contrasts demonstrated that concussion recovery was related to the increases in the LIFG and LOFC activation for 2-back, but not 1-back condition. Specifically, adolescents with concussion had significantly lower activation at baseline compared to 6-month follow-up (LIFG: t(895.01)=-4.3, p<0.001; LOFC: t(895.05)=-6.4, p<0.001) and compared to HC (LIFG: t(79.45)=-3.3, p=0.004; LOFC: t(94.06)=-3.3, p=0.004).

In the LIFG, all groups showed increased activation for 2-back vs. 1-back (HC: t(895.01)= -8.39, CONC subacute: t(895.01)= -4.47, CONC at 6-month: t(895.01)= -10.43, all p-values<0.001). In the LOFC, HC showed no difference between 2- and 1-back (t(895.05)=0.45, p=0.66), CONC subacute showed lower activation for 2-back vs. 1-back (t(895.05)= -2.7,p=0.008), while CONC at 6-month showed greater activation for 2-back vs. 1-back (t(895.05)= 3.2,p=0.001)).

To examine the relationship between brain activation and behavioral measures of working memory, we conducted mixed effects analysis with group, n-back condition, and brain activation as predictors and RT and accuracy as outcome variables. These analyses revealed significant group—by—LIFG (F(2, 845.98)=10.2, p<0.001) and group—by—LOFC (F(2, 925.07)=9.5, p<0.001) interaction effects on RT (Figure 4). Post-hoc analysis showed that these effects were driven by brain activation in the CONC group at baseline (i.e., <10 days post-concussion)) for the 2-back condition. Specifically, at baseline, the CONC with lower activation in the LIFG (slope: t(961.06)= -3.07, p=0.002) and LOFC (slope: t(911.05)= -2.97, p=0.003) also had slower RT in the 2-back condition. Interestingly, at 6-month follow-up, the same participants showed an opposite pattern of results: CONC with lower LOFC activation showed faster RT in the 2-back condition.

**Figure 4.**
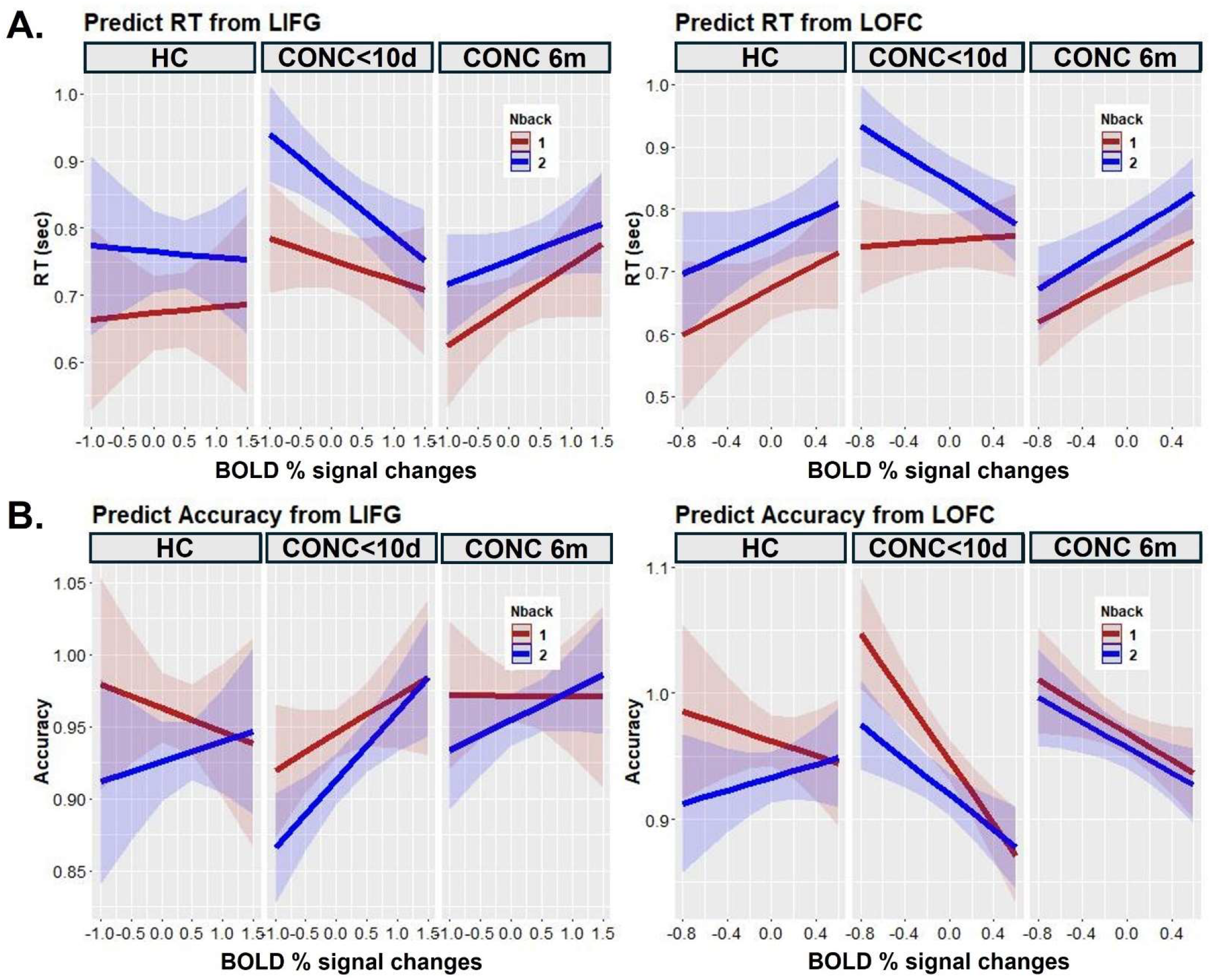
The effect of brain activation, n-back condition, and group on RT and accuracy. “CONC<10d” - adolescents with concussion at baseline, “CONC 6m” - adolescents with concussion at 6-month follow-up. “HC”-healthy controls.

There was a significant group—by—LOFC (F(2, 798.98)=4.3, p<0.05) interaction effect on accuracy. Greater activation in the LOFC corresponded to lower accuracy in concussed, but not control, adolescents at both baseline and 6-month scans (slope for CONC subacute: t(953.84)= -4.68, p<0.001; slope for CONC at 6-month: t(952.89)= -2.72, p=0.007).

## 4 DISCUSSION

Understanding behavioral and neural underpinnings of post-concussion recovery of working memory performance is critically important for improving clinical outcomes and informing better return-to-activity decisions. In this study, we examined how behavioral and neural correlates of working memory changed between the subacute period (<10 days) and 6 months of post-concussion recovery in affected adolescents. Consistent with multiple previous studies [12–16,40], participants were slower and less accurate for a more difficult 2-back vs. a less difficult 1-back condition. concussed adolescents were slower and less accurate during the subacute recovery period than at 6 months after concussion, which aligned with some longitudinal studies of concussion [11] but not the others [26]. During the subacute period, concussed adolescents were also slower than HC, which parallels previous findings [16,24].

The fMRI analysis revealed that the subacute post-concussion period was characterized by aberrant brain responses in the left inferior frontal gyrus (LIFG) and orbitofrontal cortex (LOFC). These results are largely consistent with those reported previously [27]. Specifically, affected adolescents were unable to increase activation in the LIFG for the 2-back vs. 1-back condition immediately after concussion but were able to do so after 6 months of recovery. Moreover, the magnitude of the LIFG activation changes from 1-back to 2-back did not differ for concussed adolescents at the 6-month follow-up and their non-concussed counterparts. The LIFG is a part of the working memory circuitry as per the NeuroSynth meta-analysis [41]. It plays a role in cognitive control, resolution of proactive interference [42], and updating information in working memory [43]. In our study, the inability to activate the LIFG for the 2-back condition was associated with slower RT and lower accuracy in concussed adolescents in the subacute period. Our previous resting-state functional connectivity study indicated that unlike HC showing connectivity between the IFG and dorsal attention network, concussed adolescents demonstrated no such connectivity [44]. Disconnection of the IFG from the attentional network may serve to avoid a ‘chain reaction’ and, consequently, more massive cognitive impairments when the IFG becomes dysfunctional.

The LOFC is the region located outside the working memory circuitry. The levels of activation in the LOFC were not different for 1-back and 2-back in HC, thus confirming that this region is usually insensitive to working memory load. Interestingly, in concussed adolescents, activation in this region depended on the post-concussion recovery phase. While activation associated with the 1-back condition remained the same, activation for 2-back significantly increased during 6 months of recovery. These findings may be indicative of the compensatory strategy that engages additional brain regions to perform working memory tasks after a concussion. The compensatory strategies could be related to an increase in neural effort to maintain cognitive functioning even if it goes for a price of a higher level of fatigue [45]. Engaging additional regions could help avoid excessive fatigue by distributing processing across more regions than is usually necessary. Considering that the lateral OFC is implicated in decision-making [46] and top-down attentional control and performance monitoring [47], engaging the LOFC later in the post-concussion recovery process might help to maintain a high level of behavioral performance. These findings suggest that the consequences of concussion on the brain may last beyond the point of return-to-play clearance, thus warranting a need for longer prospective studies.

Despite previous notions of emotional lability and reduced emotion regulation after concussion [48], we found no effect of emotional distractors on either behavioral or neural correlates of working memory in concussed adolescents neither immediately after the injury nor 6 months after that. This lack of findings suggests that concussion-related impairments in emotion processing and regulation are too subtle to be captured by fMRI.

One limitation of this study is the lack of longitudinal data for HC, which makes it impossible to tease apart the effect of task repetition from the effect of post-concussion recovery. Adolescents with concussion were scanned twice, so their 6-month improvement in working memory might be partially associated with task repetition. Some studies report that the task repetition effect exists even 6 months after the first exposure to the task [49]. One piece of evidence for a task repetition effect comes from the finding of higher accuracy for concussed adolescents at 6 months relative to HC at baseline. On the other hand, all participants showed very high accuracy (>90%) on the n-back task, so the variations in the accuracy are not necessarily meaningful.

Scanning HC at a 6-month follow-up would help us understand if performing the task for the second time makes the task easier or alters brain activation for difficult vs. easy tasks. The second limitation is that this study had only two scans, which did not allow us to examine the trajectory of concussion recovery. The third limitation is a rather long period between the scans. By the 6-month follow-up, all adolescents with Concussions were asymptomatic and have been cleared to return to sports and other activities. Future studies should consider scanning affected adolescents and their healthy counterparts more frequently to uncover the trajectory of post-concussion recovery. To better understand the compensatory mechanisms discussed above, future prospective studies should have a longer duration than 6 months post-injury to understand when and how brain function returns to normal.

In summary, our study provides insight into working memory-related behavioral and brain activation changes occurring across six months following concussion in adolescents. Participants experience significant improvements in RT and accuracy, as well as normalization of brain responses to the task difficulty in the LIFG and LOFC during the n-back task across a 6-month post-concussion time period. We believe that the patterns of LOFC activation reported in this study might reflect compensatory strategies to distribute neural processing and reduce neural fatigue following concussion. Considering the complexity of concussion recovery, extended prospective studies with more frequent fMRI assessments are needed to fully elucidate the dynamics of brain function recovery that may underlie behavioral performance improvements and clinical recovery in adolescents following concussion.

## Data Availability

All data produced in the present study are available upon reasonable request to the authors

## ACKNOWLEDGMENTS

The authors thank participants for taking part in this research study.

## Supplementary Materials

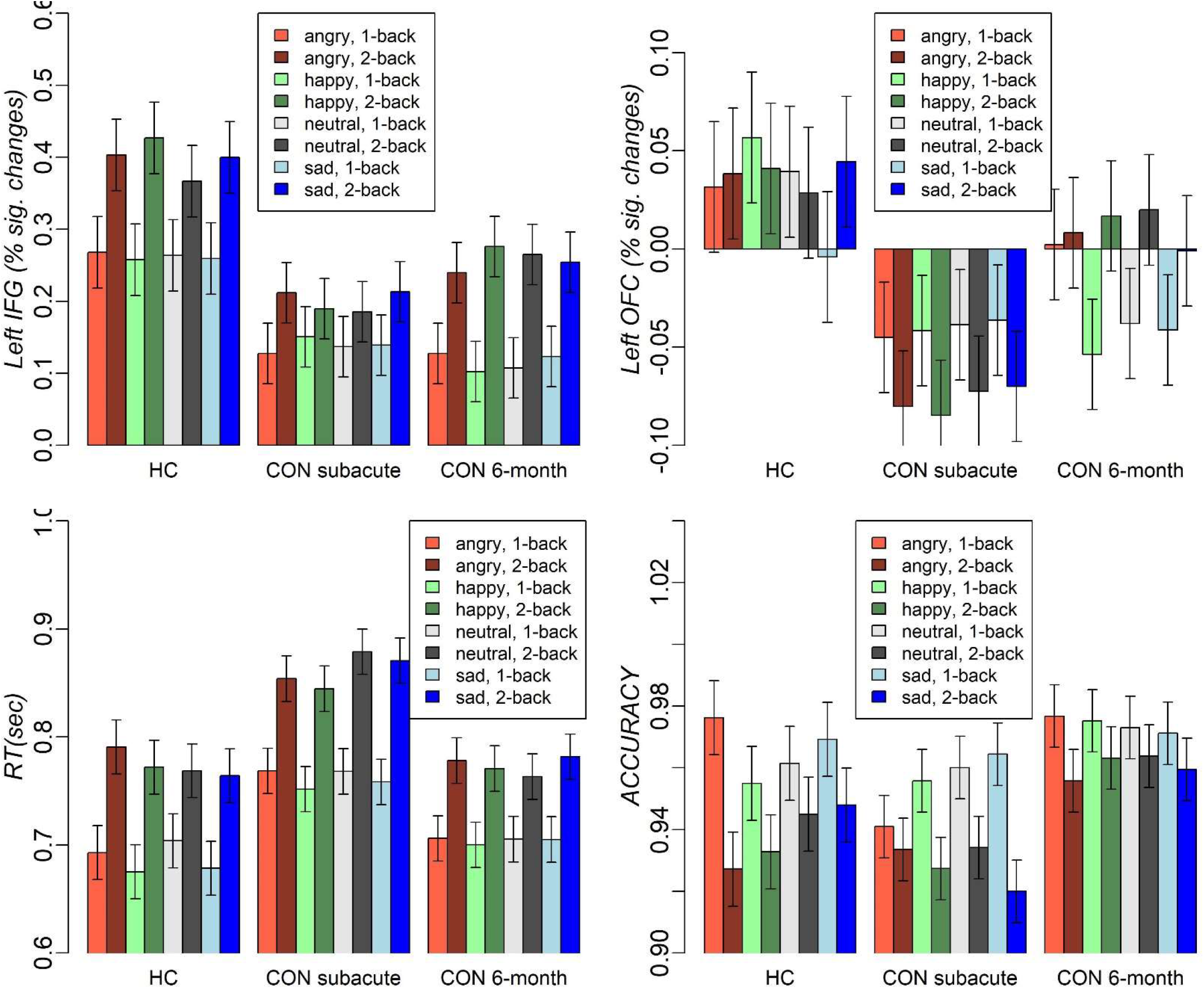

